# The general impact of haploinsufficiency on brain connectivity underlies the pleiotropic effect of neuropsychiatric CNVs

**DOI:** 10.1101/2020.03.18.20038505

**Authors:** Clara Moreau, Guillaume Huguet, Sebastian Urchs, Elise Douard, Hanad Sharmarke, Pierre Orban, Aurélie Labbe, Claudia Modenato, Sandra Martin-Brevet, Kumar Kuldeep, Charles-Olivier Martin, Khadije Jizi, Nadine Younis, Petra Tamer, Jean-Louis Martineau, Ana Isabel Silva, Aia E. Jønch, Amy Lin, Simons VIP Foundation, Jeremy Hall, Marianne B.M. van den Bree, Michael J. Owen, David E. J. Linden, Anne. M. Maillard, Sarah Lippé, Celia Greenwood, Carrie E. Bearden, Paul M. Thompson, Pierre Bellec, Sebastien Jacquemont

## Abstract

Copy number variants (CNVs) are among the most highly penetrant genetic risk factors for neuropsychiatric disorders. Their impact on brain connectivity remains mostly unstudied. Because they confer risk for overlapping conditions, we hypothesized that they may converge on shared connectivity patterns.

We performed connectome-wide analyses using resting-state functional MRI data from 436 carriers of neuropsychiatric CNVs at the 1q21.1, 15q11.2, 16p11.2, 22q11.2 loci, 4 “neutral effect” CNVs, 66 carriers of scarcer neuropsychiatric CNVs, 756 individuals with idiopathic autism spectrum disorder (ASD), schizophrenia, attention deficit hyperactivity disorder, and 5,377 controls. Neuropsychiatric CNVs showed global shifts of mean connectivity. The effect size of CNVs on relative connectivity (adjusted for the mean) was correlated with the known level of neuropsychiatric risk conferred by CNVs. Individuals with idiopathic schizophrenia and ASD had similarities in connectivity with neuropsychiatric CNVs. We reported a linear relationship between connectivity and intolerance to haploinsufficiency measured for all genes encompassed by CNVs across 18 loci. This profile involved the thalamus, the basal ganglia, somatomotor and frontoparietal networks and was correlated with lower general intelligence and higher autism severity scores. An exploratory factor analysis confirmed the contribution of these regions to three latent components shared across CNVs and neuropsychiatric disorders.

We posit that deleting genes intolerant to haploinsufficiency reorganize connectivity along general dimensions irrespective of where deletions occur in the genome. This haploinsufficiency brain signature opens new avenues to understand polygenicity in psychiatric conditions and the pleiotropic effect of CNVs on cognition and risk for neuropsychiatric disorders.

**One sentence summary:** Neuropsychiatric CNVs across the genome reorganize brain connectivity architecture along dominant patterns contributing to complex idiopathic conditions.

## Introduction

Genomic copy number variants (CNVs) are deletions (DEL) or duplications (DUP) of more than 1000 base pairs of DNA. Rare CNVs with large effects have been associated with a range of neurodevelopmental and psychiatric conditions *(1, 2)*. Twelve recurrent CNVs have been individually associated with autism spectrum disorder (ASD) *(3)*, eight with schizophrenia (SZ) *(4)*, and eight with attention deficit hyperactivity disorder (ADHD) *(5)* but studies have shown that ultra-rare CNVs at many more genomic are also associated with these conditions *(4, 6)*.

Functional connectivity (FC) studies have provided critical insight into the architecture of brain networks involved in neuropsychiatric disorders (NPs), but only a few studies have investigated networks modulated by CNVs *(7–9)*. These large effect-size mutations can shed light on pathways connecting genetic risk to brain endophenotypes, such as FC. In a previous study, we characterized the connectome-wide effects of four CNVs that confer high risk for NPs (NP-CNVs). Deletions and duplications at the 16p11.2 and, to a lesser extent, at the 22q11.2 locus were associated with mirror effects on global FC *(9)*. For 16p11.2 deletion carriers, overconnectivity predominantly involved the ventral attention, motor, and frontoparietal networks relative to controls. 22q11.2 deletion carriers showed global underconnectivity, involving the anterior and lateral default mode network (DMN) and the limbic network. Connectivity profiles of the thalamus, somatomotor, posterior insula and cingulate showed significant similarities between NP-CNVs and idiopathic ASD, SZ but not ADHD.

Previous studies were mainly performed one mutation at a time, with the notion that the function of genes would shed light on the relationship between molecular mechanisms and phenotypes. Results from this approach have raised several questions including 1) How specific are the effects of CNVs, and are there any general rules linking the gene content of CNVs to intermediate brain phenotypes? 2) How can one pursue the study of rare variants beyond the handful of CNVs and single nucleotide variants (SNV) frequent enough to conduct an individual association study? The hypothesis of shared neuroimaging alterations across NP-CNVs was recently investigated in 21 carriers of CNVs across the 22q11.2, 15q11.2, 1q21.1, 16p11.2 or 17q12 loci *(10)*. Analysis of diffusion-weighted imaging (DWI) measures from the cingulum bundles suggested that macro- and microstructural properties were associated with the level of risk for psychiatric disorders conferred by CNVs. Using T1-weighted data, Warland and colleagues showed that the volumes of three subcortical brain regions (thalamus, hippocampus, and nucleus accumbens) were significantly reduced in a group of 49 carriers of 4 SZ-associated CNVs in the UK Biobank (16p11.2 duplication, 22q11.2 deletion, 15q11.2 deletion, and 1q21.1 deletion) *(11)*.

In the current study, we asked two questions: 1) Do previous observations of connectivity alterations associated with 16p11.2 and 22q11.2 extend to other genomic loci? and 2) How can we close the gap between the exponentially expanding landscape of rare neuropsychiatric variants and the knowledge of their effects on intermediate brain phenotypes? We recently tackled a similar question by investigating the statistical relationship between the coding gene content of rare CNVs and their effect on intelligence quotient (IQ). We showed that around 75% of the effect-size of any CNV on IQ can be explained by linear models using the sum of the “probability of being loss-of-function intolerant” (pLI) scores *(12)* of all genes encompassed in the CNV *(13)*. The pLI (probability loss-of-function intolerant) score is the probability that a given gene is intolerant to “protein loss of function” (pLoF). The score measures selective pressure and is based on lower-than-expected rates of variants leading to haploinsufficiency in the general population *(12)*.

We aimed to 1) characterize the connectivity-profiles of CNVs at genomic loci previously associated with neurodevelopmental disorders (1q21.1, 15q11.2, 16p11.2 and 22q11.2), 2) assess whether connectivity-profiles of NP-CNVs may represent dimensions observed in idiopathic ASD, SZ, or ADHD, and 3) investigate the relationship between measures of pLI and connectivity across genomic loci. We gathered rsfMRI data on 502 carriers of deletions or duplications at the 1q21.1, 2q13, 15q11.2 (BP1-BP2), 15q13.3, 16p11.2 proximal, and 22q11.2 genomic loci as well as 66 carriers of scarcer NP-CNVs at 8 additional genomic loci *(6)*. Among these carriers were included 4 “neutral effect” CNVs without prior association to neuropsychiatric conditions (2q13 CNVs, 15q13.3 and TAR-1q21.1 duplications) *(4, 6, 14)*. Three out of the five genetic-first cohorts used in this study have not yet been published. We also analyzed 756 subjects with idiopathic ASD, schizophrenia, or ADHD, and 5,377 controls (Table 1).

**Table 1.** CNV carriers, individuals with idiopathic psychiatric conditions and controls after MRI quality control. Chr: chromosome number, coordinates are presented in Megabases (Mb) according to Hg19. DEL: deletion; DUP: duplication; IPCs: Idiopathic Psychiatric Conditions; SZ: schizophrenia, ASD: Autism Spectrum Disorder; ADHD: Attention-Deficit/Hyperactivity-Disorder n = tot /clin: total number of participants /number of participants clinically ascertained. Age (in years); M: male; Motion: framewise displacement (in mm). Quantitative variables are expressed as the mean ± standard deviation. All sites scanned controls and sensitivity analyses were performed to investigate the potential bias introduced by differences in site, age and sex. Odd-ratios (OR) for the enrichment of CNVs in ASD and schizophrenia were previously published (a *(6)*, b *(4)*). OR for the enrichment of CNVs in ADHD were not available. The four ‘Neutral-effect CNVs’ are highlighted by a light grey background. Additional NP-CNVs (n=66) included in the pLI analysis include NRXN1 (n=2 deletion), 13q12.1 (n=5 deletion, n=2 duplication), 16p12.1 (n=1 deletion, n=3 duplication), 16p13.11 (n=4 deletion, n=6 duplication), 17p12 (n=5 deletion; n=1 duplication), TAR (n=2 deletion), 22q11.2 [B-D] (n=3 deletion, n=23 duplication), 2q11.2 (n=1 deletion, n=2 duplication), 16p11.2 distal (n=1 duplication), 7q11.23 distal (n=1 duplication), 15q13.3 (n=1 deletion), 14q32 (n=1 deletion) and 2 carriers of multiples CNVs. Detailed information relative to diagnosis, IQ, and motion, are available in Supplementary Tables 2-3

| CNV (hg19) | Status | n tot /clin | Age | Sex (M) | Motion | Sites | OR ASD | OR SZ |
| --- | --- | --- | --- | --- | --- | --- | --- | --- |
| **15q11.2**<br>chr15:<br>22.81-23.09 | DEL | 66 / 1 | 63 (7.6) | 42% | 0.18 (0.06) | 3 | n.s | 2 (b) |
|  | DUP | 71 / 1 | 62.5 (6.9) | 45% | 0.18 (0.06) | 3 | n.s. (a) | n.s. (a) |
| **15q13.3**<br>31.08-32.46 | DUP | 40 / 0 | 62.6 (6.7) | 37% | 0.18 (0.05) | 2 | n.s. (a) | n.s. (a) |
| **2q13**<br>chr2:<br>110.86-110.98 | DEL | 36 / 0 | 63.3 (7.3) | 31% | 0.18 (0.06) | 2 | - | - |
|  | DUP | 30 / 0 | 62.8 (7) | 40% | 0.17 (0.05) | 2 | - | - |
| **1q21.1**<br>chr1:<br>146.53-147.39 | DEL | 25 / 9 | 42.4 (20.4) | 56% | 0.18 (0.06) | 7 | 3 (a) | 4 (b) |
|  | DUP | 16 / 7 | 42.36 (21) | 31% | 0.2 (0.06) | 7 | 5 (a) | 4 (b) |
| 145.39-147.39 | DUP-TAR | 18 / 0 | 58.6 (7.9) | 56% | 0.17 (0.05) | 2 | - | - |
| **22q11.2**<br>chr 22:<br>19.04-21.47 | DEL | 43 / 43 | 17 (7) | 55% | 0.18 (0.07) | 1 | 32 (a) | 68 (b) |
|  | DUP | 19 / 11 | 36 (23.1) | 42% | 0.18 (0.09) | 4 | n.s. (a) | 0.15 (b) |
| **16p11.2**<br>chr 16:<br>29.65-30.20 | DEL | 41 / 37 | 18.8 (19) | 58% | 0.22 (0.08) | 6 | 14 (a) | 12 (a) |
|  | DUP | 31 / 29 | 30.5 (17) | 64% | 0.21 (0.09) | 4 | 14 (a) | 9 (b) |
| Additional CNVs for pLI analysis |  | 66 / 12 | 57 (16) | 45% | 0.18 (0.06) | 5 | - | - |
| **CNVs Controls** |  | 4427 / 0 | 61.31 (11) | 47% | 0.19 (0.06) | 7 | - | - |
| **Idiopathic Psychiatric Conditions** | **SZ** | 242 / 242 | 33.6 (9.24) | 73% | 0.16 (0.06) | 9 | - | - |
|  | **ASD** | 225 / 225 | 16 (6.5) | 100% | 0.18 (0.05) | 12 | - | - |
|  | **ADHD** | 289 / 289 | 11.5 (2.8) | 78% | 0.15 (0.04) | 6 | - | - |
| **IPCs Controls** |  | 950 / 0 | 18.8 (10) | 70% | 0.15 (0.04) | 27 | - | - |

## Results

### Neuropsychiatric CNVs cause global shifts of functional connectivity

Deletions and duplications of several genomic loci showed a shift in mean FC (Figure 1a-e). There was a positive association between the number of genomic copies (deletion=1, duplication=3) and global connectivity for the 22q11.2 and 1q21.1 CNVs, and negative gene dosage effect for the 16p11.2 CNVs (Figure 1a, 1d-e, Supplementary Tables 4-5). A negative shift was observed for both 15q11.2 and 2q13 duplications (Figure 1b, 1c). In all subsequent analyses, we investigated relative FC, which is computed by adjusting for mean whole-brain connectivity (mC-adjusted).

**Figure 1.**
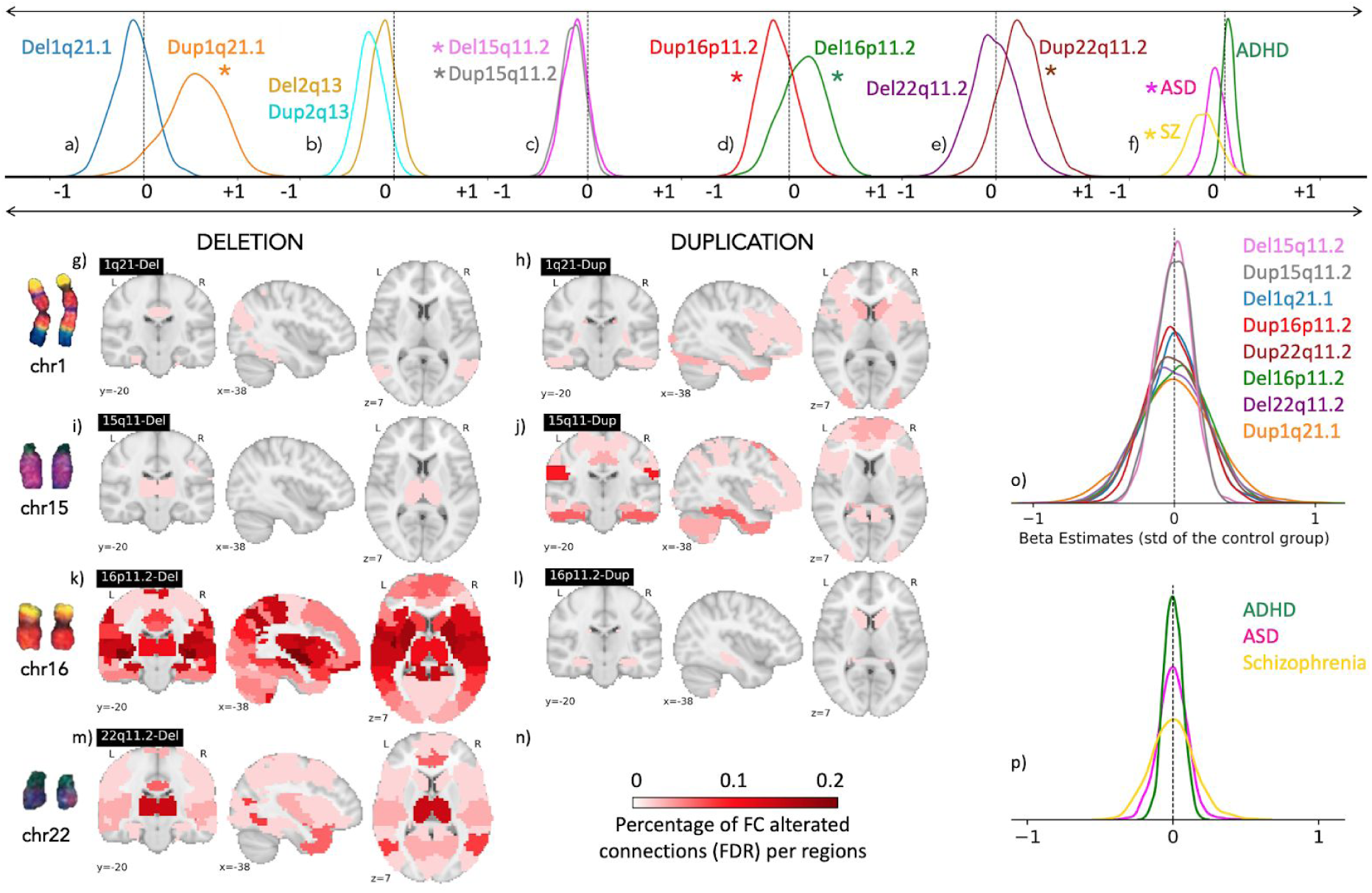
(a-e) Global effects of CNVs on connectivity. Density plots represent the distribution of the 2,080 beta estimates for the connectome wide association study (CWAS, whole-brain contrast of cases versus controls) of the CNVs, SZ, ASD, ADHD groups. Stars represent a significant global shift in mean FC. The same density plots are shown after adjusting for mean connectivity (o-p). X-axis values of all density plots represent z-scores of the Beta estimates, which were obtained from linear models computed using z-scored connectomes based on the variance of the control group. Brain maps (g-n) represent the percentage of altered connections (FDR corrected) per region. Del=Deletion, Dup=Duplication, ASD: autism spectrum disorder; SZ: schizophrenia; ADHD: attention deficit hyperactivity disorder; chr=chromosome.

### CNV severity is linked to the effect-size on relative connectivity

We previously showed that the severity of a CNV’s impact on cognition is strongly associated with measures of intolerance to haploinsufficiency such as the pLI *(13)*. We, therefore, tested the relationship between pLI scores and the size of a CNV’s effect on FC across the 7 genomic loci (listed in Table 2). We showed a significant correlation between pLI scores and effect sizes of deletions (*r*=0.89, *p*=0.03). There was no significant relationship for duplications.

**Table 2.** The number of significantly altered connections (FDR corrected) for each connectome wide association study (n=15) after adjusting for mean connectivity (Supplementary Table 4-5-6). ∑ pLI: sum of pLI of all genes encompassed in each CNVs. pLI: the **p**robability of being **L**oss of function **I**ntolerant is a measure of gene’s intolerance to haploinsufficiency. DEL: deletion; DUP: duplication; ASD: autism spectrum disorder; SZ: schizophrenia; ADHD: attention deficit hyperactivity disorder. min-max: minimum-maximum of z-scored beta values; var: variance of z-scored beta values; n pos: number of positive connections; n neg: number of negative connections.

| Loci / $\sum$ pLI | CNV | n pos | n neg | min-max | var |
| --- | --- | --- | --- | --- | --- |
| 1q21.1<br>1.7 | DEL | 0 | 3 | [-1; 0.7] | 0.04 |
|  | DUP | 9 | 4 | [-1; 1] | 0.08 |
| TAR (3.4) | DUP | 0 | 0 | / | 0.04 |
| 2q13<br>0 | DEL | 0 | 0 | / | 0.02 |
|  | DUP | 0 | 0 | / | 0.02 |
| 15q11.2<br>1.7 | DEL | 1 | 0 | [-0.4; 0.5] | 0.02 |
|  | DUP | 11 | 16 | [-0.5; 0.4] | 0.02 |
| 15q13.3 (1.6) | DUP | 0 | 0 | / | 0.02 |
| 16p11.2<br>7.06 | DEL | 76 | 84 | [-0.9; 1.4] | 0.06 |
|  | DUP | 1 | 3 | [-0.9; 0.7] | 0.04 |
| 22q11.2<br>10.5 | DEL | 25 | 21 | [-0.9; 0.8] | 0.06 |
|  | DUP | 0 | 0 | [-0.7; 0.8] | 0.05 |
| ADHD |  | 0 | 0 | / | 0.01 |
| ASD |  | 21 | 9 | [-0.3; 0.4] | 0.01 |
| Schizophrenia |  | 203 | 233 | [-0.4; 0.6] | 0.02 |

### Relative regional connectivity is robustly altered by high-risk neuropsychiatric CNVs

The 16p11.2 deletion significantly altered 160 connections (76 positives, 84 negatives, FDR, q<0.05) with beta values ranging from -0.8 to 1.4 z-scores (z-scores based on the variance of the control group, Table 2, Figure 1.k, Supplementary table 6). The altered connections mostly involved the ventral and dorsal posterior insula, the pre-supplementary motor cortex, the putamen, dorsal precuneus, and the left inferior parietal lobule. The 16p11.2 duplication significantly altered 4 connections (1 positive, and 3 negatives), with beta values ranging from -0.9 to 0.7 z-scores). The altered connections mostly involved the amygdala-hippocampus complex, the cerebellum crus-II and VIIIab, and the caudate and accumbens nuclei (Table 2, Figure 1.l, Supplementary Table 6).

The 22q11.2 deletion was associated with over-connectivity in 25 connections and underconnectivity in 21 connections (FDR, q < 0.05), with beta values ranging from -0.95 to 0.8 z-scores. The regions showing the strongest FC alterations included the thalamus, the dorsal anterior and posterior cingulate cortices, the lateral fusiform gyrus, the temporal pole, and the anterior insula (Table 2, Figure 1.m, Supplementary Table 6). The 22q11.2 duplication did not show FC alterations that survived FDR.

### 15q11.2 and the 1q21.1 CNVs have mild effects on relative connectivity

The 15q11.2 deletion was associated with overconnectivity of one connection (FDR, q < 0.05) between the thalamus and the ventrolateral somatomotor network (Figure 1i, Table 2, Supplementary Table 6). In the 15q11.2 duplication carrier group, 27 connections were significantly altered (16 negatives, and 11 positives, beta values [-0.51; 0.36], Table 2). Altered connections primarily involved the supramarginal, inferior temporal, occipitotemporal gyri, and the temporal pole (Figure 1.j, Supplementary Table 6).

The 1q21.1 deletion was associated with underconnectivity of three connections (FDR, q < 0.05) between by the lateral fusiform gyrus, the dorsal precuneus, the lateral occipitotemporal gyrus, the dorsal visual stream, and the dorsal posterior cingulate cortex, with beta values ranging from -1.0 to 0.67 z-scores, (Table 2, Supplementary Table 6, and Figure 1.g). The 1q21.1 duplication showed 13 connections that were significantly altered (4 negatives, and 9 positives, FDR, q < 0.05), with beta values ranging from -0.98 to 1.0 z-scores. Altered connections mostly involved the caudate nucleus, the posterior lateral visual network, the temporal pole, the cerebellum Crus-I, and the putamen (Figure 1.h, Table 2, Supplementary Table 6).

We did not detect any significant effects of the 4 neutral effect CNVs (Tar 1q21.1 duplication, 15q13.3 duplication and 2q13 deletion and duplication) on connectivity (Supplementary Table 6).

### Neuropsychiatric CNVs and idiopathic psychiatric conditions show whole-brain FC similarities

We compared (Mann-Whitney) spatial similarities between CNV FC-profiles and IPC, with spatial similarities between CNV FC-profiles and controls (Figure 2c-d). This was performed for all 12 CNVs and 3 IPCs (Figure 3a). Out of the 36 correlations, 12 survived FDR. Of those, most were observed between large effect-size neuropsychiatric CNVs, SZ and ASD. Neutral CNVs (2q13 CNVs and 15q13.3 and TAR-1q21.1 duplication) did not show similarities with FC-profiles of individuals with IPC.

**Figure 2.**
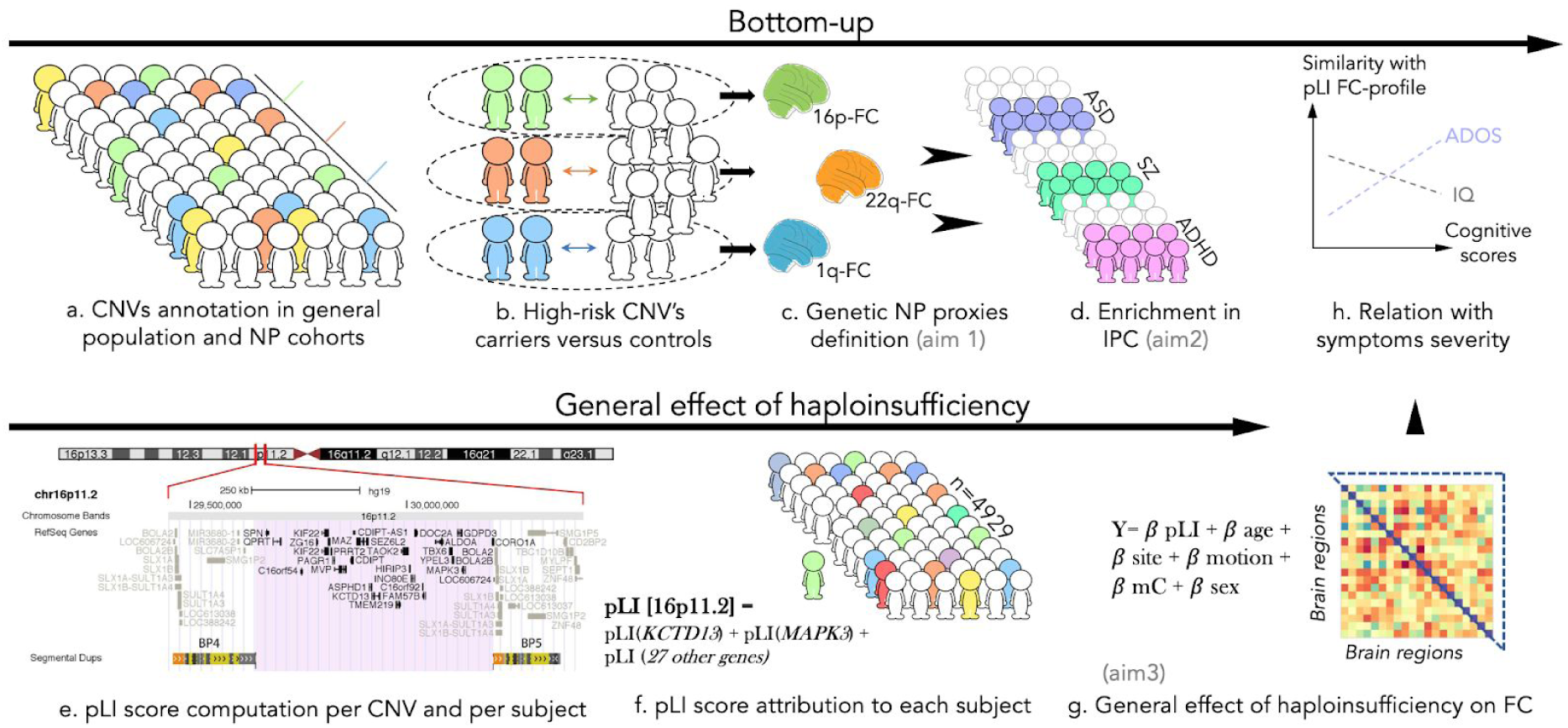
Method overview: a) CNVs are identified in unselected and neuropsychiatric populations. b) Connectome wide association studies (CWAS) between CNVs carriers and controls define a CNV-FC-profile (c) for each CNV (aim 1). d) Comparing (Mann-Whitney) 1] the spatial similarity between CNV FC-profile and individuals with IPCs, and 2] the spatial similarity between CNV FC-profile and controls (aim 2). e) To investigate the general effect of pLI on FC, we used CNVs at 18 genomic loci. We computed the sum of pLI of all genes encompassed within each genomic loci. f) Each carrier obtains a score corresponding to the gene content of his CNV. g) Mass univariate linear models draw a relationship between the pLI score (X) of each individual, and each of the 2080 functional connections (Y) h) Correlation at the regional level between pLI FC-profile and symptoms severity scores. NP: neuropsychiatric conditions; 16p: 16p11.2; 22q: 22q11.2; 1q:1q21.1; ASD: autism spectrum disorder; SZ: schizophrenia; ADHD: attention deficit hyperactivity disorder; mC: mean-connectivity adjustment.

**Figure 3.**
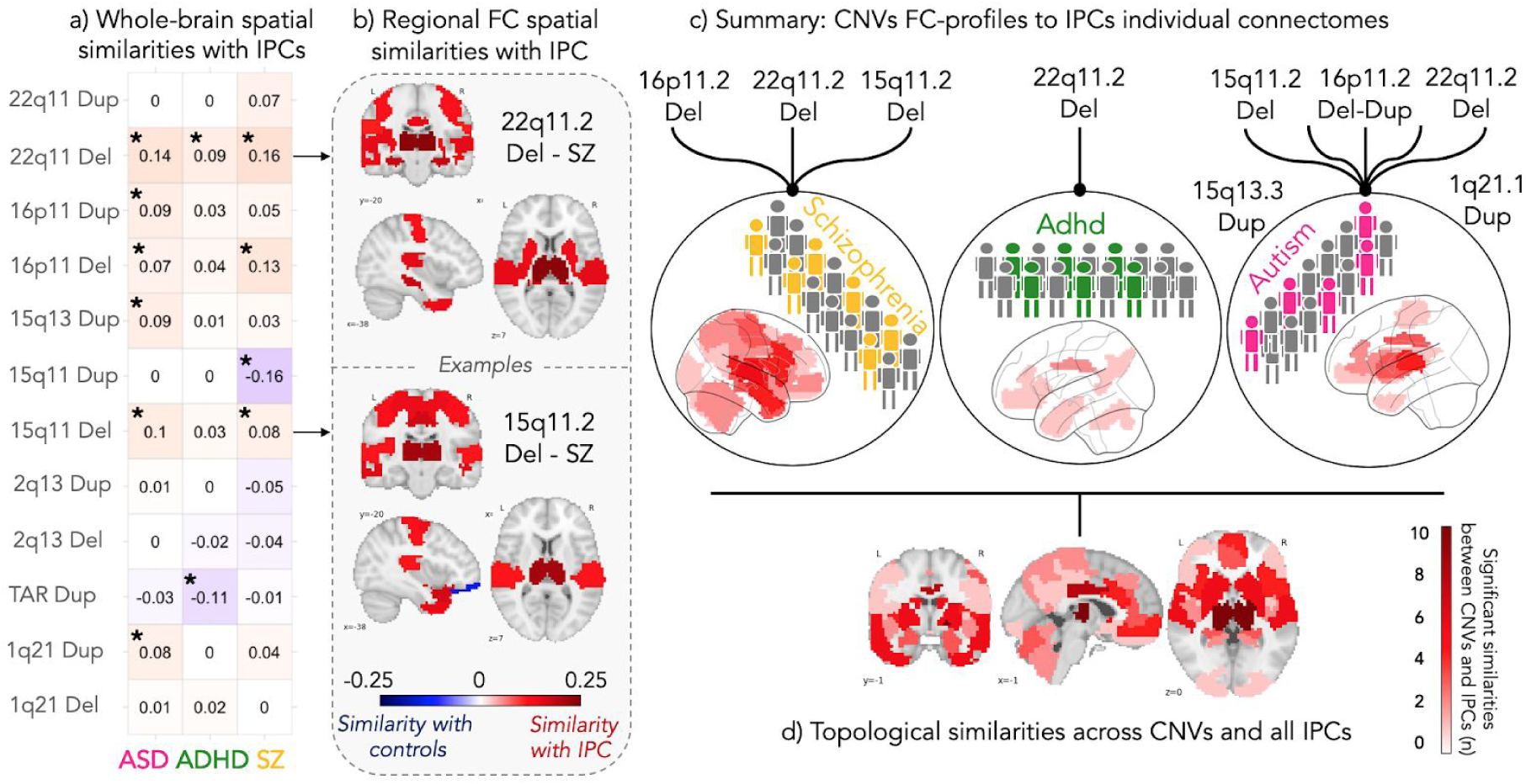
Spatial similarities between FC-profiles of CNVs and idiopathic psychiatric conditions at the regional and connectome-wide level. (a) The whole-brain FC-profiles of CNVs were correlated to the individual FC-profiles of subjects with a psychiatric diagnosis, and their respective controls. The effect size (Mann-Whitney test statistic, rank biserial correlation) of the spatial similarity between the CNVs-FC-profiles and individuals with IPC are detailed in the table. Positive (red) reflects higher similarity between CNVs-FC-profiles and individuals with IPC, while negative (blue) reflects higher similarity with controls. Stars represent significant similarities after FDR correction for 36 tests. (b) The whole-brain FC-profiles of CNVs were decomposed into 64 seed-based regions and compared to the individual FC-profiles of subjects with a psychiatric diagnosis, and their respective controls. Regions with higher similarities between CNVs and IPC are presented in red hues for 2 examples: 15q11.2 deletion-FC-profile (left side) and 22q11.2 deletion-FC-profile (right side) to individual connectomes of SZ and controls. Colors reflect the effect size (rank-biserial correlation) of the similarity between CNV and idiopathic patients (all the 36 brain maps are available in Supplementary Figure 3, with corresponding statistical values in Supplementary Table 7). (c) The spatial similarity between the whole-brain FC-profiles of CNVs and connectomes of IPC individuals. Connectors represent significant similarities between group-level CNVs-FC-profiles and the individual connectomes of either IPC cases or controls. Brain maps summarize FC similarities at the regional level between CNVs and ASD (right), ADHD (middle) and SZ. The color scale represents the number of times a region shows significant similarities between CNV FC-profiles and IPCs. (d) Brain map summarizes the frequency of regions involved in FC similarities between all NP-CNVs and all IPCs. Eg. the thalamus showed significant similarities in 9 comparisons:15q11.2 deletion and ASD; 15q11.2 deletion and SZ; 16p11.2 deletion and ASD; 16p11.2 deletion and SZ; 22q11.2 deletion and ASD, 22q11.2 deletion and SZ; 1q21.1 duplication and SZ; 15q13.3 duplication and ASD; 22q11.2 duplication and SZ. ASD: autism spectrum disorder; SZ: schizophrenia; ADHD: attention deficit hyperactivity disorder; Del: deletion; Dup: duplication; 22q11: 22q11.2, 16p11: 16p11.2; 1q21: 1q21.1, TAR: 1q21.1-TAR, 15q11: 15q11.2, 15q13: 15q13.3.

### Connectivity similarities between neuropsychiatric CNVs and idiopathic psychiatric conditions involve the thalamus, the basal ganglia and the posterior cingulate cortex

We investigated whether whole-brain FC similarities between individuals with SZ, ASD, ADHD and CNVs were driven by particular regions. To do so, we applied the same approach as described above at the regional level by decomposing the FC-profiles of each CNV into 64 seed regions. We found that a set of regions including the thalamus, the caudate, the putamen, the posterior cingulate, temporal pole, and anterior insula exhibited high degrees of similarity between all neuropsychiatric CNVs FC-profiles and individuals with IPC (figure 3b, red regions, Supplementary Table 7, and Supplementary Figure 3). Only a few regional-level CNVs FC-profiles showed higher similarities with controls (Figure 3b, blue regions, Supplementary Figure 3). Individuals with SZ and ASD demonstrated the highest level of similarity with all neuropsychiatric CNVs compared to their respective controls. We did not detect significant similarities between the FC-profiles of “neutral” CNVs (2q13 deletion and duplication, and TAR-1q21.1 duplication) and individuals with IPC (Figure 3, Supplemental Figure 3, Supplementary Table 7). None of the similarity was correlated with motion.

### Haploinsufficiency is associated with a profile of dysconnectivity shared across genomic loci

To investigate the potential general effects of haploinsufficiency on FC, we used a model previously developed to estimate the effect size of any CNVs on general cognitive abilities *(13)*. This model used as an explanatory variable the sum of pLI scores of all genes encompassed in all deletions and duplications carried by an individual to explain FC (Figure 2g) *(12)*. We analyzed all CNVs available: 502 CNVs carriers at 18 genomic loci and 4,427 individuals who did not carry a detectable CNV and thus had a pLI score of zero (Table 1, Figure 2g, Supplementary methods).

The pLI-associated profile for deletions (hereinafter referred to as haploinsufficiency profile) was characterized by a higher FC in 60 connections mainly involving the thalamus and a lower FC in 58 connections in the anterior cingulate, the pre-supplementary motor area, and the dorsomedial prefrontal cortex (Figure 4a-b, Supplementary Table 6). Since this linear model may be influenced by CNVs with the largest pLI scores, we performed a sensitivity analysis and showed that removing the 16p11.2, and the 22q11.2 deletions did not substantially impact the observed pattern of dysconnectivity (haploinsufficiency profile before and after exclusion were correlated at r=0.77, r=0.72, respectively). Our results suggest that this haploinsufficiency FC profile is present across genomic loci with an effect size correlated to pLI. We were likely underpowered to detect a pLI-associated FC-profile for duplications (only 3 connections survived FDR).

**Figure 4.**
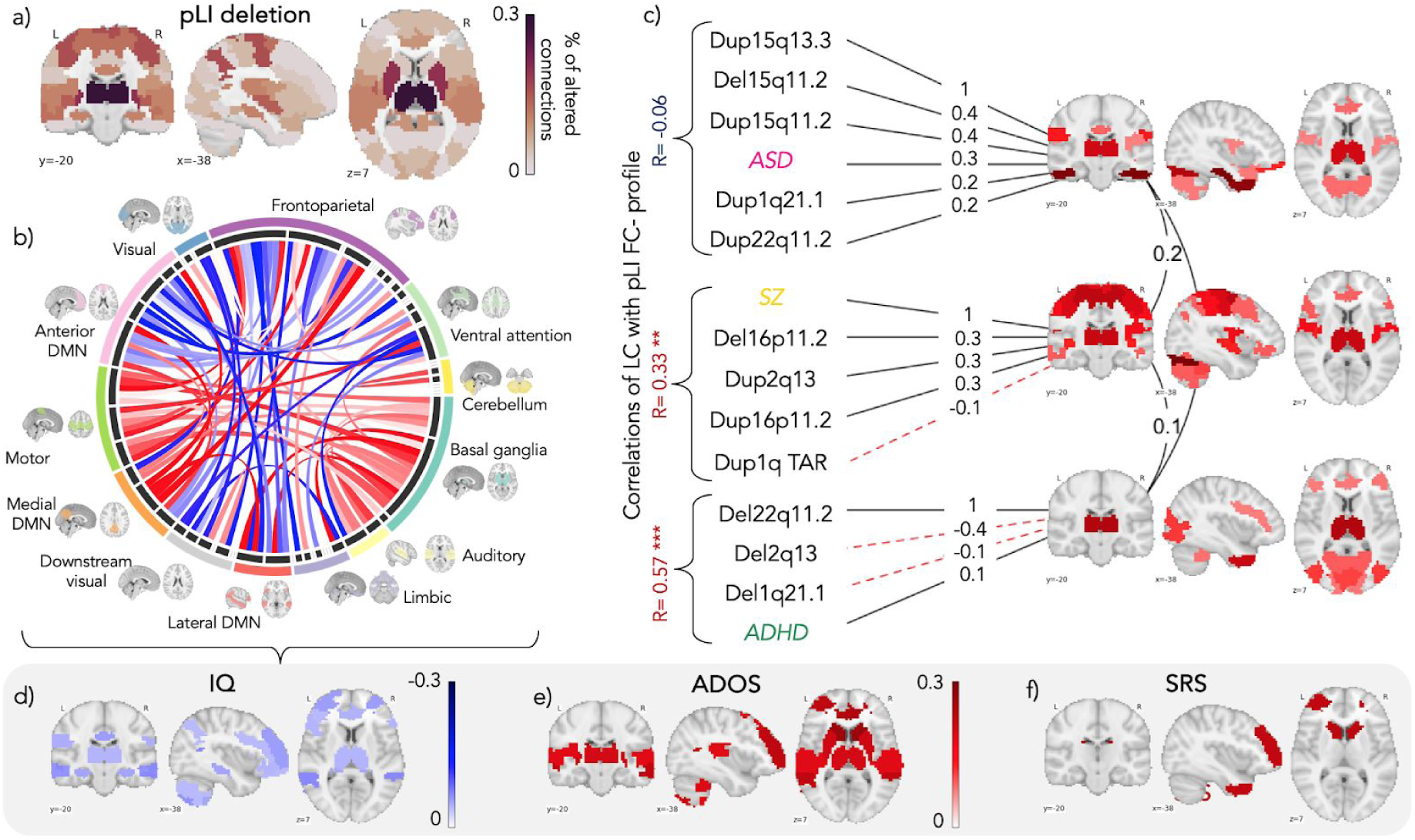
(a-b) Relationship between dysconnectivity and haploinsufficiency measured by pLI across genomic loci. (a) Brain map represents the percentage of altered connections (FDR corrected) per region. (b) Each chord represents a functional connection between 2 regions that are significantly associated with intolerance to haploinsufficiency (measured by pLI). All 64 seed regions are represented in the dark grey inner circle of the chord diagram. The width of the seed region in the grey inner circle corresponds to the number of altered connections. Seed regions are grouped into 12 functional networks (outer ring). Networks are represented in 12 brains around the diagram. Red chords represent overconnectivity and blue chords underconnectivity. Full names are available in Supplementary Table 10. (c) Exploratory Factor Analysis results across 15-FC profiles of alterations converging on 3 latent components (LCs). Value on each arrow represents standardized EFA loading (see Supplementary Method). We reported the 64 values for the corresponding 3 LCs into the 3 brain maps (red: positive contribution, blue: negative contribution, Supplementary Table 9). (d) Individuals with higher similarity to the haploinsufficiency FC-profile have lower measures of general intelligence and higher autism severity measures. Colour codes represent Pearson r computed between the 64 regions of the haploinsufficiency FC-profile and the regional profiles of individuals based on their cognitive and behavioural measures. IQ: Intelligence Quotient, ADOS: Autism Diagnostic Observation Schedule, SRS: Social Responsiveness Scale.

We investigated the relationship between the haploinsufficiency FC-profile and general intelligence, autism, schizophrenia and ADHD severity measures (Figure 2.h). Individuals with lower general intelligence scores showed similarity with 20 out of the 64 regions of the haploinsufficiency FC-profile (Figure 4d-f, Supplementary Table 8). This negative association was significant across all 3 cohorts with general intelligence measures (UKBB non-carriers, ABIDE autism and controls, ADHD cases and controls). Autism severity scores (ADOS and SRS) also increased in individuals with higher similarities to pLI-FC-profile in 11 and 3 regions respectively (Supplementary Table 8). Regions driving correlation with intelligence and autism measures were partially overlapping (figure 4d). None of these similarities was correlated to motion.

### A parsimonious set of FC dimensions may underlie the dysconnectivity observed across CNVs and idiopathic psychiatric conditions

We asked if dysconnectivity profiles across all CNVs and IPCs could be summarized by latent components (LCs). We used the FC-profiles of 12 CNVs delineated in aim 1 (those with sample size allowing CWAS) and 3 IPCs obtained by CWAS (Figure 1o-p, Supplementary results). We performed an exploratory factor analysis (EFA) using maximum likelihood as a fitting procedure across all CNVs and IPC FC-profiles. The EFA identified 3 LCs that explained 28% of the variance between FC-profiles (Tucker-Lewis Index (TLI) of factoring reliability = 0.93, Root Mean Square Error Approximation index (RMSEA) =0.01, *p*=0.42 for the null hypothesis that 3 factors were sufficient). Regions contributing the most across LCs included the thalamus, the temporal pole, the anterior cingulate, and the ventromedial prefrontal cortex (Supplementary Table 9). The third LC showed a high spatial similarity with the haploinsufficiency FC-profile (*r*=0.57) (Figure 4c).

## Discussion

### Main findings

We provided the first connectome-wide characterization of CNVs at the 1q21.1 and 15q11.2 loci and conducted a systematic comparison across 8 neuropsychiatric CNVs. Deletions and duplications at 6 out 8 CNVs were associated with a global shift in mean connectivity with the largest effect at the 1q21.1 duplication locus. The effect size of CNVs on relative FC (mean-connectivity adjusted) was correlated with the known level of NP-risk conferred by CNVs. We identified connectivity architecture similarities between high-risk NP-CNVs and individuals with ASD, SZ and to a lesser extent ADHD. These similarities were driven by the thalamus, the posterior cingulate cortex, and the anterior insula. Four neutral control CNVs exhibited far less similarities with IPCs than large effect size NP-CNVs. Intolerance to haploinsufficiency measured by pLI score was related to a FC profile characterized by increased connectivity in the thalamus, basal ganglia, the somatomotor network and medial DMN, and decreased connectivity in the limbic, frontoparietal and anterior DMN. This haploinsufficiency profile was associated with lower measures of intelligence across three cohorts and an increase in autism severity scores. Using exploratory factor analysis, we validated these regions by showing their contribution to three latent components explaining 28% of FC variance across CNVs and neuropsychiatric conditions.

### Bottom-up genetic-first versus top-down approaches

The effect sizes of rare variants on functional neuroimaging traits are concordant with effects previously measured for the same variants on brain structure, cognitive and behavioural traits *(13, 15)*. This is in striking contrast with neuroimaging studies of behaviorally defined groups of patients that have required very large samples to reach reproducible results. On average, the effect sizes observed for FC and brain structure in SZ, ASD and ADHD range from 0.3 to 0.15 and lower *(16, 17)* which is discordant with the severity of those conditions that lie well beyond 2 standard-deviation with respect to their impact on behavioural and adaptive traits. Therefore, while brain intermediate endophenotypes remain elusive in heterogeneous psychiatric conditions, genetic first strategies have the potential to provide key insights into underlying biological mechanisms.

### Pleiotropic effects in neuropsychiatric CNVs on brain connectivity

GWAS studies have shown that the same set of SNPs confer risk for a range of different conditions, suggesting pleiotropic effects *(18)*. Similarly, CNVs increase risk for a range of psychiatric disorders including ASD, SZ, ADHD, as well as obsessive-compulsive, oppositional defiant, and tic disorders *(19)*. Mechanisms underlying these apparent pleiotropic effects are unclear. Recent systematic cross-CNV analyses reported that impairments in many behavioural and cognitive traits were broadly similar for 12 NP-CNVs *(19)*. There were only moderate qualitative profile differences in cognitive and behavioral measures between NP-CNVs*(19)*. Consistent with these findings, we showed that CNVs present similarities with several IPCs at the connectivity level. This may be related to connectivity-dimensions such as the FC profile defined by intolerance to haploinsufficiency associated with increased risk across several psychiatric conditions and cognitive deficits *(13, 20)*. In line with these observations, cross-psychiatric diagnoses studies have delineated FC dimensions very similar to those associated with haploinsufficiency and involving the somatosensory motor network, DMN, thalamus and subcortical structures *(21)*. This is consistent with our current and previous findings showing that the FC patterns shared between CNVs and IPC were correlated to IQ and SRS*(9)*.

### Haploinsufficiency reorganizes brain connectivity according to a general FC-profile

We extended to connectivity some models which were initially developed to estimate the effect size of CNVs on IQ *(13)*. This approach showed that deleting genes intolerant to haploinsufficiency (measured by pLI) may lead to a pattern of dysconnectivity irrespective of where the deletion occurs in the genome. This haploinsufficiency profile was associated with a decrease in intelligence measures in the general population and disease cohorts and increased severity on autism assessments. Such findings may help decipher why the same linear model using pLI as an explanatory variable can explain 75% of the effect of deletions on IQ irrespective of genomic location and point to mechanisms explaining why 70%–100% of any 1-MB windows in the human genome contributes to increased risk for SZ and ASD *(20, 22)*. Intolerance to haploinsufficiency measured by pLI reflects a negative selection pressure unrelated to a particular molecular function. Non-specific effects of haploinsufficiency on cognition, behavior and FC could be related to emerging properties of the genome, rather than a limited set of biological pathways *(23)*. In other words, changing gene dosage at any node of the genomic network may alter its efficiency leading to a measurable effect on brain organization and behavior.

### The Thalamus is a central hub across psychiatric disorders and rare neuropsychiatric variants

Recent models of thalamic functions revealed a far more complex contribution than a simple passive relay with extensive connections to the entire cerebral cortex. Functional MRI studies have demonstrated that thalamocortical and corticothalamic pathways are engaged in memory, attention, and mental representations through specific thalamic subdivisions *(24, 25)*. fMRI studies have reported involvement of the thalamus across several NPs including ASD, SZ, and major depression *(26–28)*. Our study also highlighted the thalamus as a functional hub highly sensitive to altered gene dosage across loci. The thalamic connectivity-pattern in high-risk NP-CNVs carriers also showed the highest similarities with those of ASD, SZ, and ADHD. Finer parcellation of the thalamus *(24)* will provide insight into the relationship between functional alterations related to CNVs and IPCs.

## Limitations

Previous studies have shown that the effect size of duplications is 2 to 3 fold smaller than deletions for cognitive *(20)* and neuroanatomical measures *(15, 29)*. A similar phenomenon was observed in this study and much larger samples of duplication carriers will be required to accurately characterize their effects on FC.

Our study was designed to search for shared effects but we are not implying that most of the FC alterations are common across CNVs. It will require much more data on many more genomic loci to delineate the potential specific effects of any given genomic variant.

This multisite study associating clinically and non-clinically ascertained cohorts may have introduced biases. Several confounding factors including sex bias, age differences, and medication status may also have influenced some of the results. However, carefully conducted sensitivity analyses, investigating all of these confounders, and performing sensitivity analyses matching control group for sex, site, age, motion and excluding IDP with medications provided similar results. Finally, we could not perform EFA with all 2,080 connections due to the insufficient number of CNV carriers. By moving to regional patterns, directionality is no longer taken into account. Larger samples are required to confirm and further characterise these latent components.

## Conclusion

Our results suggest that deletions and duplications across the genome may converge upon a parsimonious set of connectivity dimensions involved in cognition and idiopathic NPs. Such findings raise the question on the nature of the relationship between the molecular functions of genes included in CNVs and functional changes of large scale brain networks. General haploinsufficiency connectivity profiles will likely extend to a large number of genomic loci and may help decipher why broad groups of genomic variants confer risk to the same neuropsychiatric conditions.

## Materials and Methods

### Sample

We analyzed 6,635 individuals from nine datasets (Table 1, Supplementary Materials and Methods).

#### CNVs carriers and controls

Genetic-first cohorts were recruited based on the presence of a CNV, regardless of symptomatology, by five consortia (three out of five have never been published before): the Simons Variation in Individuals Project (VIP) consortium data (16p11.2 and 1q21.1 CNVs carriers) *(30)*, the University of California, Los Angeles (22q11.2 CNVs carriers), the Brain Canada cross NP-CNVs project (CHU Sainte Justine, Montreal, Canada), the Define cross NP-CNVs Project (Cardiff, UK), and the Lausanne Prisma project (16p11.2 and 1q21.1 CNVs carriers) (see Supplementary Materials and Methods for individual dataset description).

CNVs were also identified in an unselected population (UK Biobank) (see Supplementary Materials and Methods for the CNV calling procedure and final sample description).

#### Idiopathic psychiatric conditions and respective controls

Individuals with idiopathic ASD and their respective controls were sampled from the ABIDE1 multicenter dataset *(31)*. Individuals with idiopathic SZ and their respective controls were obtained from aggregated fMRI data of 10 studies. Individuals diagnosed with ADHD (DSM-IV) and their respective controls were obtained from the ADHD-200 dataset *(32, 33)*(see Supplementary Materials and Methods for individual dataset description).

Imaging data were acquired with site-specific MRI sequences. Each cohort analyzed in this study was approved by the research ethics review boards of the respective institutions. Signed informed consent was obtained from all participants or their legal guardian before participation. Secondary analyses of the listed datasets for the purpose of this project were approved by the research ethics review board at Sainte Justine Hospital. After data preprocessing and quality control, we included a total of 6,635 individuals (Table 1).

### Preprocessing and QC procedures

All datasets were preprocessed using the same parameters with the same Neuroimaging Analysis Kit (NIAK) version 0.12.4, an Octave-based open-source processing and analysis pipeline *(34)*. Preprocessed data were visually controlled for quality of the co-registration, head motion, and related artefacts by two raters (Supplementary Materials and Methods).

### Computing connectomes

We segmented the brain into 64 functional seed-based regions defined by the multi-resolution MIST brain parcellation *(35)*. FC was computed as the temporal pairwise Pearson’s correlation between the average time series of the 64 seed-based regions, and then Fisher-z transformed. The connectome of each individual encompassed 2,080 connectivity values: (63×64)/2 = 2016 region-to-region connectivity + 64 within seed-based region connectivity. We chose the 64 parcel atlas of the multi-resolution MIST parcellation as it falls within the range of network resolution previously identified to be maximally sensitive to functional connectivity alterations in neurodevelopmental disorders such as ASD (Supplementary Table 10) *(36)*. We corrected for multiple comparisons using a false discovery rate strategy *(37)*.

Statistical analyses were performed in Python using the scikit-learn library *(38)*. Analyses were visualized in Python and R. Code for all analyses and visualizations is available online through the GitHub platform with Jupyter notebook: https://github.com/claramoreau9/NeuropsychiatricCNVs_Connectivity.

### Statistical analyses

All of the following analyses are summarised in Supplemental Materials and Methods (Objective and methods, Supplementary Table 1).

#### Connectome-wide association studies (CWAS)

We performed fifteen CWAS: comparing FC between cases and controls for five CNVs (15q11.2, 1q21.1, 2q13, 16p11.2 and 22q11.2, for deletion and duplication carriers), for TAR-1q21.1 and 15q13.3 duplications carriers, and for three idiopathic psychiatric cohorts (ASD, SZ, and ADHD). Controls were pooled across all CNV cohorts (n=4,427). Controls were separately pooled across the three idiopathic groups (IPCs, n=950, Table 1). FC was standardized (z-scored) based on the variance of the respective control group. CWAS was conducted by linear regression at the connectome level, in which z-scored FC was the dependent variable and clinical status the explanatory variable. Models were adjusted for sex, scanning site, head motion, mean connectivity, and age. We determined whether a connection was significantly altered by the clinical status effect by testing whether the β value (regression coefficient associated with the clinical status variable) was significantly different from 0 using a two-tailed *t*-test. This regression test was applied independently to each of the 2,080 functional connections. We corrected for the number of tests (2,080) using the Benjamini-Hochberg correction for FDR at a threshold of *q* < 0.05 *(37)*, following the recommendations of Bellec *et al*. 2015 *(39)*.

We defined the global FC shift as the average of the β values across all 2,080 connections. We tested whether the observed global FC shifts were significantly different from zero by conducting a permutation test, shuffling the clinical status labels of the individuals included in each CWAS (using 10,000 replications). We thus estimated a valid permutation-based *p*-value associated with the observed global FC shift *(40)*.

Two additional CWAS were performed to assess the linear effect of pLI deletion and duplication scores on FC. The CNV pLI annotation is described in the Supplementary Materials and Methods. This analysis was performed only among the CNVs cohorts, controlling for sex, scanning site, head motion, mean connectivity, and age. FC was standardized (z-scored) based on the variance of the entire sample.

#### Similarity of whole-brain connectivity-profiles between idiopathic psychiatric conditions and CNVs

We tested the similarity between dysconnectivity measured across the 3 IPCs and the 12 CNVs. This similarity was tested by correlating, at the whole-brain level, individual connectomes of cases and controls of IPCs to the CNVs-connectivity-profiles (group level; Figure 2cd). Controls used to compute the similarity with psychiatric conditions have not been used to compute the CNVs-FC profiles in the first place. The group-level FC-profile was defined as the 2,080 β values obtained from the contrast of cases vs. controls (aim1.2). This was repeated between all CNVs (n=12) and the 3 conditions (n=36 similarity tests).

Individual connectomes of IPC cases and controls were used after independently adjusting for sex, scanning site, mean connectivity, head motion, and age. Similarity scores were derived by computing Pearson’s correlations between the whole brain connectomes. We asked whether IPC cases compared to the IPC controls had significantly higher (or lower) similarity to whole-brain CNV-profile using a Mann-Whitney U test. We reported significant group differences after FDR correction accounting for the 36 tests (*q* < 0.05).

#### Similarity of regional connectivity-profiles between idiopathic conditions and CNVs

The same approach described above was performed at the regional level. We calculated a similarity score between individual adjusted connectomes and the 12 CNVs FC-profiles. FC-profiles were broken down into 64 region-level FC-profiles and similarity scores were derived by computing Pearson’s correlations between the 64 β values associated with a particular region. For each region, we tested whether individuals with a psychiatric diagnosis had significantly higher (or lower) similarity to CNVs FC-profiles than controls using a Mann-Whitney U test. We reported significant group differences after FDR correction (*q* < 0.05) for the number of regions (n=64).

We investigated the relationship between severity scores and similarity with pLI-FC profile. Similarities of individuals with pLI FC-profiles were correlated (Pearson’s r) with severity scores. The p-values associated with these correlations were corrected for multiple comparisons (FDR, *q* < 0.05).

#### Exploratory factorial analysis

Exploratory Factor Analysis (EFA) was performed using the maximum likelihood (mle) method. Factors were allowed to rotate. Analyses were performed using the *psych* and the *stats* packages in R 3.4.1 *(41)*. Factor models were fit iteratively and compared using three criteria: TLI ≥ 0.90, RMSEA ≤ 0.10 and a smaller Bayesian Information Criteria relative to other models. The model with the best fit has been retained. We used FC-profiles of 12 CNVs and 3 IPCs obtained by CWAS (aim 1.2). The EFA identified 3 latent components (LC) to obtain a non-significant p-value (test for the null hypothesis that 3 factors were sufficient). We extracted standardized EFA loading scores per LC. We used the Nilearn package *(38)* to report the 64 standardized loading scores per LC into 3 brain maps. We computed the Pearson correlation between the pLI deletion FC-profile and each of the three LCs.

## Supplementary Material, Methods, and Results

## Data Availability

Processed connectomes are available through a request to the corresponding authors.

https://github.com/claramoreau9/NeuropsychiatricCNVs_Connectivity

## Author contributions

C.M., S.J., and P. B. designed the overall study and drafted the manuscript.

### Analyses

C.M. and S.U. processed 90% of all the fMRI data and performed all imaging analyses.

G.H., J-L. M., and E. D. performed the CNVs calling and pLI annotations

P.O. preprocessed the SZ data.

A.L. contributed to the computation and the interpretation of the EFA data

H.S. performed 3/4 of the UKBiobank fMRI preprocessing.

AE.J., C.M., K.K, C.G., and S.M-B. contributed to the interpretation of the data and reviewed the manuscript.

### Data collection

D.L. and A.D.S. provided the Cardiff Define CNV fMRI data.

S.L., K.J, C.O., P.T., N.Y. recruited/scanned patients for the Brain Canada Montreal CNV project.

C.E.B., A.L. provided the UCLA 22q.11.2 fMRI data.

A.M. provided the Lausanne CHUV fMRI data.

D.L., M.O., M. V.d.B., J.H, and A.I.S. provided the Cardiff CNV fMRI data

The Simons Variation in Individuals Project Consortium provided the 16p11.2 data.

All authors provided feedback on the manuscript.

### Data and materials availability

Processed connectomes are available through request to the corresponding authors.

